# Comparison of machine learning methods for clinical data imputation among a real-world lung cancer cohort

**DOI:** 10.1101/2022.06.12.22276306

**Authors:** Daniel X. Yang, Yongfeng Hui, Betsy Wang, Arman Avesta, Charlotte Zuber, Henry S. Park, Sanjay Aneja

## Abstract

**PURPOSE:** Cancer registries are important sources of real-world data (RWD) that reveal insights into practice patterns and cancer patient outcomes, but the prevalence of missing data can be high. Machine learning (ML) imputation methods can be applied to large RWD sets, but the performance of these approaches within cancer registries is unclear.

**METHODS:** We identified non-small cell lung cancer (NSCLC) patients within the National Cancer Database diagnosed in 2014 with complete data in 19 variables of known clinical and prognostic significance. We generated synthetic missing data for each variable, then performed imputation using substitution (control) and five different ML approaches. Imputation efficacy was measured by normalized root-mean-square error (RMSE) for continuous variables and proportion of falsely classified entries (PFC) for categorical variables. We also measured algorithm runtimes and the impact of incorporating imputed values on survival modeling.

**RESULTS:** 50,790 NSCLC patients were included for this study, with 81 features for each patient after data preprocessing. Among the tested ML methods, SoftImpute had the lowest RMSE (best performance) for continuous variables ranging from 0.071 to 0.080 for 10% to 50% missing data, and MissForest had the lowest PFC (best performance) for categorical variables ranging from 0.251 to 0.311 for 10 to 50% missing data. SoftImpute had a runtime of 3.28×10^−4^ seconds per patient record, and MissForest averaged 2.96×10^−3^ seconds per patient record. Deep learning imputation using a denoising autoencoder did not achieve improved performance despite higher algorithm runtimes. Cox models incorporating ML imputed data achieved similar C-index ranging from 0.787 to 0.801 for all ML methods tested.

**CONCLUSION:** ML imputation achieved promising performance for NSCLC patients within a large national cancer registry.

## Introduction

Cancer registries are important sources of real-world data (RWD) that have generated insights spanning cancer epidemiology, practice patterns, and patient outcomes.^1,2^ Missing data represents one of the more significant limitations when comparing registry data to data collected in protocoled clinical studies. Missing data may occur only in part due to random chance and can be a surrogate for missing clinical information that was not documented in the medical record. Among non-small cell lung cancer (NSCLC) patients within a national cancer database, patients with missing data were more likely have advanced stage.^3^ Importantly, when compared to patients of the same stage, those with missing registry data demonstrated worse outcomes suggesting that excluding such patients in registry analysis may lead to biased findings.

Data imputation is one possible solution to increase the representation of patients with missing data within registry analyses. Traditional imputation methods have demonstrated mixed results in terms of accuracy and reducing potential bias, and the selection of appropriate imputation models remains challenging for clinical researchers.^4,5^ Recently, there is increasing interest in using machine learning (ML) based imputation methods which have demonstrated promising results on diverse types of datasets.^6,7^ However, the efficacy of these imputation techniques has not been well studied in cancer registry data, and ML imputation approaches are rarely incorporated into observational studies within oncology.

In this study, we compare the efficacy of ML methods to impute missing data on a national registry of NSCLC patients. Specifically, we evaluate the performance of ML approaches for missing data imputation and how cancer patient survival models are affected when incorporating imputed clinical information.

## Methods

### Data Source

We identified non-small cell lung cancer (NSCLC) patients with complete data in 19 variables (Supplemental Table 1) with known clinical and prognostic significance within the National Cancer Database (NCDB).^8–14^ In this study, we focused on NSCLC given our previous work suggesting missing data may be used to identify patients with worse associated survival outcomes. We chose patients diagnosed in 2014 to allow sufficient follow up to examine downstream overall survival, and patient records with complete data were chosen given a reference value is needed to compare the performance. Categorical variables with a large number of categories were recoded to a fewer number of clinically relevant categories consistent with prior clinical studies.^15^

### Imputation Methods

Imputation techniques inherently rely on learning underlying representations within non-missing aspects of the dataset. Five imputation methods were studied based on previously shown efficacy on diverse datasets and differences in underlying algorithms. Machine learning based imputation methods were compared to simple substitution methods where the mean was used for continuous variables and the mode was used for categorical variables.

#### k Nearest Neighbor (KNN)

A KNN method imputes missing values for a patient using a weighted average of neighborhood of similar patients. The weighted average is calculated using the mean squared difference on features for which two patients have complete data.

#### Iterative Single Value Decomposition (SVD)

The iterative SVD method is an implementation of the imputation approach described by *Troyanskaya et al* that uses single value decomposition to obtain a set of mutually orthogonal patterns of non-missing variables which can be linearly combined to approximate all variables in the dataset.^16^

#### SoftImpute

The SoftImpute method described by *Mazumder et al* similarly uses single value decomposition for dataset completion, but is potentially more computationally efficient than IterativeSVD by regularizing using the nuclear norm of the dataset.^17^

#### MissForest

The MissForest method described by *Stekhoven et al* leverages multiple decision trees to learn the dataset representation and predict missing values. This method has been shown to be effective in mixed datasets with both continuous and categorical variables.^18^

#### Multiple Imputation with Denoising Autoencoders (MIDAS)

The Multiple Imputation with Denoising Autoencoders described by *Lall et al* is an unsupervised deep learning method.^19^ MIDAS trains denoising auto-encoders to reconstruct the original dataset after introducing additional missingness. The trained auto-encoders learn representations of the data to impute missing values seen in the original dataset.

### Comparison of Imputation Methods

To compare imputation methods, first a “complete dataset” without any missing values was created from the NCDB. A “missing dataset” was subsequently created by randomly removing data values within a fixed proportion of records across all variables of interest. Values for outcome variables (follow up time and vital status) were not removed while creating the “missing dataset”. Imputation methods were tested by comparing imputed values within the “missing dataset” to those found in the “complete dataset”. (Figure 1)

**Figure 1.**
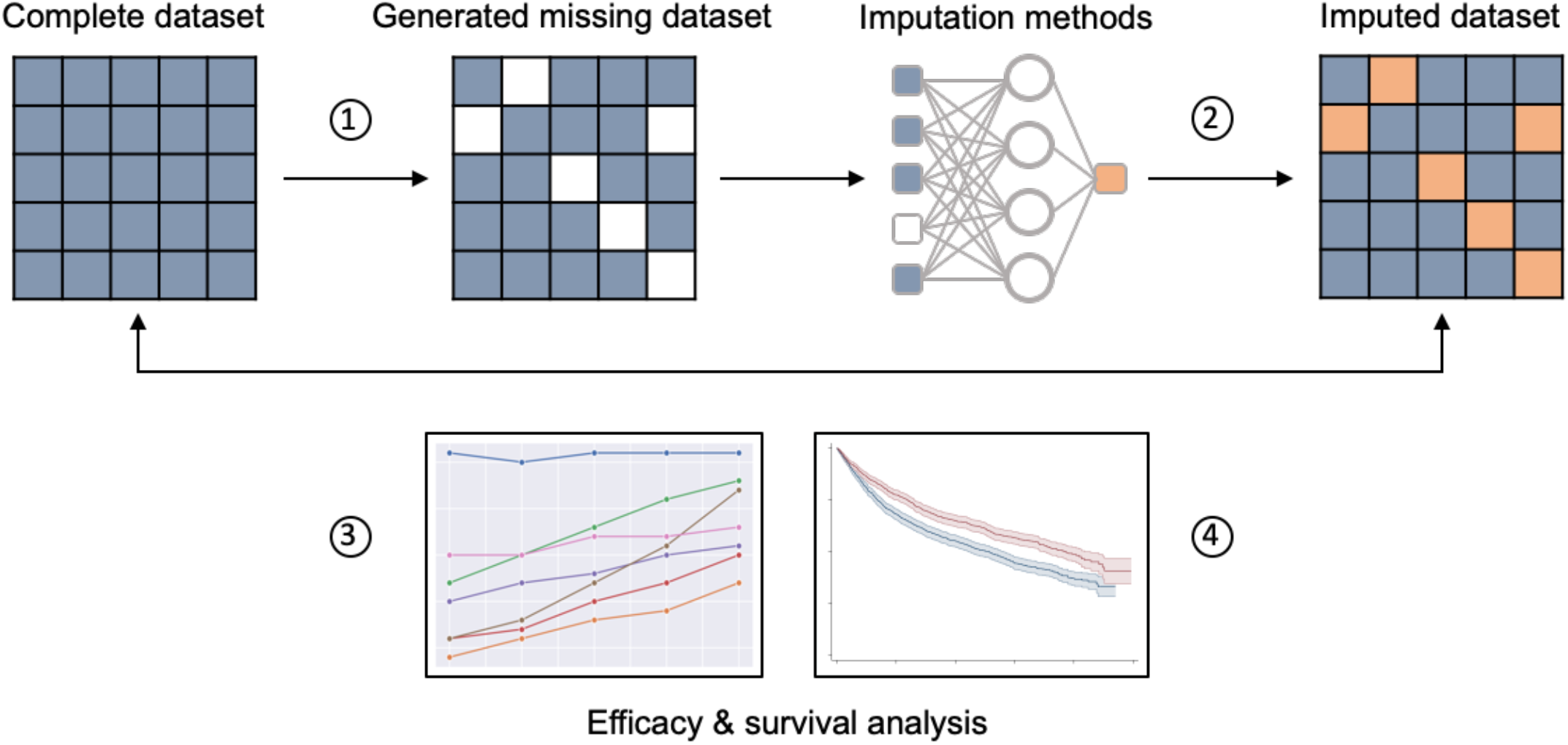
Schematic of experimental approach. From a complete dataset, we (1) generate synthetic missing values in each variable for 10-50% of patient records, then (2) apply five different machine learning approaches to impute the missing clinical information. Imputation performance is measured by (3) efficacy metrics and impact on (4) Cox survival models.

To evaluate performance across different degrees of missingness, multiple “missing datasets” were created with fixed proportions of missing data ranging from 10%-50%.

Given our previous work suggesting metastatic patients are most likely to have missing data in cancer registries, imputation methods were also compared using a “missing dataset” with a two-fold increase in proportion of missing data for patients with metastatic NSCLC compared to non-metastatic patients.^3^

### Performance of Imputation Methods

For continuous variables, imputation efficacy was measured by root-mean-square error (RMSE) between imputed values and their original counterparts. For categorical variables, imputation efficacy was measured by proportion of falsely classified entries (PFC) compared to their original counterparts. Lower RMSE and PFC values suggest a more effective imputation method. To compare RMSE across different variables, all numeric values were 0-1 scaled. PFC was calculated by summing the total number of values where the original and imputed values differed, then dividing by the total number of imputed data values.

In order to investigate how incorporation of ML imputed clinical values impacted survival modeling, Cox regression models were created using the complete “ground truth” dataset and compared to the imputed datasets. Coefficients of fitted Cox regression models were compared using mean absolute error (MAE) and mean squared error (MSE). Performance between Cox models were compared using Harrell’s concordance index (C-index).^20^

Finally, computational efficiency of different imputation methods were compared by measuring the algorithm runtimes on a high-performance computing server using 16 virtual processors and 42 gigabytes of memory. Runtimes were recorded as seconds per patient record to allow for comparison if applied to larger or smaller datasets.

#### Statistical Considerations

All experiments and analyses were performed using Python 3.7 and Stata 16. Imputation methods were implemented from open-source Python software libraries scikit-learn, fancyimpute, missingpy, and midaspy.^21–24^ The code to reproduce the results are available on our laboratory GitHub repository.^25^

## Results

### Patient characteristics

50,790 NSCLC patients diagnosed in 2014 were included in the study analysis. Patients records were included if they had complete data in the variables used in this study. Our study population was 50.2% male, 86.8% white, and 53.0% did not have recorded co-morbidities. The most common overall NSCLC stage among patients in this study was stage I (42.0%) followed by stage IV (25.2%). The median follow-up was 18.7 months (Table 1).

**Table 1.** Patient characteristics for the complete dataset used in our study.

|  |  |
| --- | --- |
|  | Total |
|  | N=50,790 |
| Age at Diagnosis | 69.0 (62.0-76.0)<sup>a</sup> |
| Sex |  |
| Male | 25,473 (50.2%)<sup>b</sup> |
| Female | 25,317 (49.8%) |
| Race |  |
| White | 44,076 (86.8%) |
| Black | 5,143 (10.1%) |
| Other | 1,571 (3.1%) |
| Charlson-Deyo Score |  |
| 0 | 26,907 (53.0%) |
| 1 | 15,877 (31.3%) |
| 2 | 5,825 (11.5%) |
| >=3 | 2,181 (4.3%) |
| NCDB Analytic Stage Group |  |
| Stage 0 | 174 (0.3%) |
| Stage I | 21,335 (42.0%) |
| Stage II | 6,979 (13.7%) |
| Stage III | 9,508 (18.7%) |
| Stage IV | 12,783 (25.2%) |
| Occult (lung only) | 11 (0.0%) |
| Last Contact or Death, Months from Dx | 18.7 (6.4-33.8) |

<sup>a</sup> Median (Interquartile Range) is reported for continuous variables
<sup>b</sup> N (%) is reported for categorical variables

### Imputation efficacy of ML methods

ML methods had improved imputation efficacy compared to substitution for both continuous and categorical variables. However, imputation efficacy decreased with increasing levels of missingness. The relative performance of ML methods also differed by variable data type. Among continuous variables, SoftImpute achieved the lowest RMSE (best performance) of 0.071 to 0.080 for 10% to 50% missing data, whereas the RMSE for mean substitution remained relatively constant between 0.090 to 0.091. Among categorical variables, MissForest achieved the lowest PFC (best performance) ranging from 0.251 to 0.311 for 10 to 50% missing data, whereas the PFC for mode substitution remained relatively constant between 0.406 to 0.407 (Figure 2).

**Figure 2.**
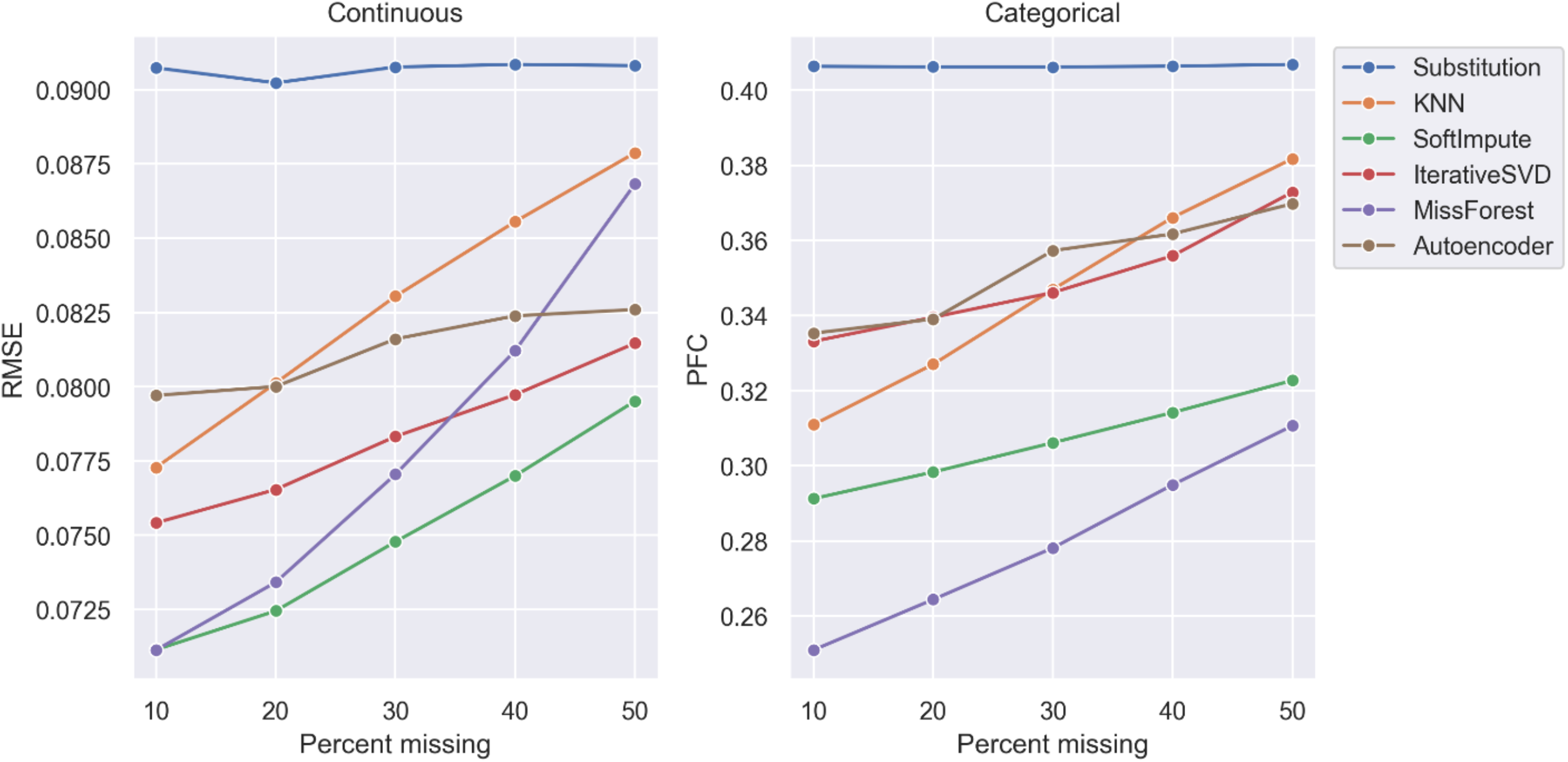
Performance of imputation methods at varying levels of missing data. Root-mean-square error (RMSE) is shown for continuous variables and proportion of falsely classified (PFC) entries is shown for categorical variables.

**Figure 3.**
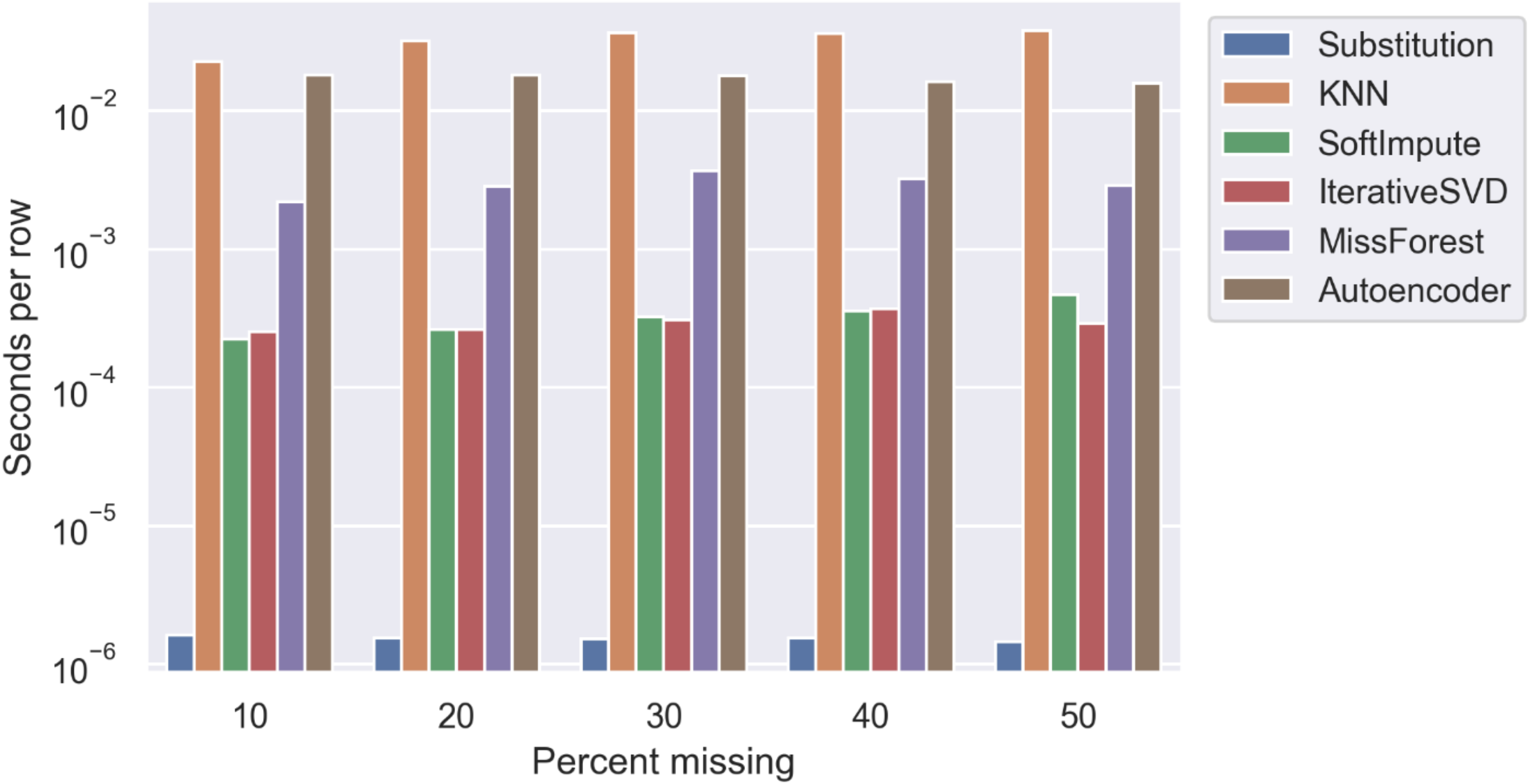
Average algorithm runtime in seconds per sample.

When missing data was introduced with a two-fold increase among metastatic NSCLC patients as compared to non-metastatic patients, similar trends in imputation efficacy were also observed. SoftImpute achieved lowest RMSE for continuous variables, and MissForest achieved lowest PFC for categorical variables (Supplemental Figure 1).

### Variable-level imputation efficacy

Variables describing tumor stage and treatment information achieved the greatest relative improvement in imputation efficacy with ML methods compared to substitution. On average, ML methods achieved a 75.0% improvement over substitution methods for imputing TNM clinical M stage, 58.4% improvement for imputing overall group stage, and 56.7% improvement for imputing type of primary site surgery. Conversely, for variables describing Race and Charlson-Deyo Score, ML methods did not outperform mode substitution (Table 2).

**Table 2.** Variable-level imputation performance at 20% missing.

|  | **Sub** | **KNN** | **SI** | **IS** | **MF** | **DA** | **Average change<sup>a</sup> (range)** |
| --- | --- | --- | --- | --- | --- | --- | --- |
| **Continuous variables, RMSE** |  |  |  |  |  |  |  |
| Age | 0.141 | 0.133 | 0.121 | 0.128 | 0.121 | 0.132 | 9.92% (5.8 - 14.3) |
| Distance to Hospital | 0.031 | 0.034 | 0.030 | 0.030 | 0.032 | 0.032 | -3.26% (-10.7 - 1.7) |
| Tumor Size | 0.041 | 0.038 | 0.035 | 0.037 | 0.037 | 0.039 | 8.23% (3.1 - 13.0) |
| Number of Lymph Nodes | 0.101 | 0.074 | 0.065 | 0.069 | 0.067 | 0.075 | 30.38% (25.5 - 35.3) |
| **Categorical variables, PFC** |  |  |  |  |  |  |  |
| Sex | 0.505 | 0.475 | 0.428 | 0.474 | 0.443 | 0.478 | 8.95% (5.4 - 15.1) |
| Insurance Status | 0.349 | 0.366 | 0.344 | 0.472 | 0.261 | 0.402 | -5.72% (-35.3 - 25.1) |
| Radiation Treatment | 0.357 | 0.235 | 0.236 | 0.268 | 0.175 | 0.252 | 34.68% (25.0 - 51.0) |
| Sequence of Radiation and Surgery | 0.060 | 0.047 | 0.058 | 0.060 | 0.021 | 0.043 | 23.37% (0.0 - 64.5) |
| Chemotherapy Treatment | 0.545 | 0.363 | 0.324 | 0.353 | 0.295 | 0.373 | 37.28% (31.4 - 45.9) |
| Sequence of Systemic Therapy and Surgery | 0.146 | 0.086 | 0.103 | 0.090 | 0.036 | 0.081 | 45.72% (29.3 - 75.0) |
| Race | 0.128 | 0.136 | 0.128 | 0.128 | 0.134 | 0.135 | -2.97% (-6.1 - 0.0) |
| Primary Site Surgery | 0.506 | 0.238 | 0.205 | 0.246 | 0.149 | 0.256 | 56.74% (49.4 - 70.5) |
| Charlson-Deyo Comorbidity Score | 0.463 | 0.514 | 0.463 | 0.544 | 0.479 | 0.535 | -9.50% (-17.5 - 0.0) |
| Tumor Grade | 0.535 | 0.534 | 0.463 | 0.554 | 0.475 | 0.555 | 3.52% (-3.8 - 13.5) |
| Stage Group | 0.579 | 0.266 | 0.221 | 0.251 | 0.170 | 0.297 | 58.38% (48.7 - 70.6) |
| Median Household Income | 0.683 | 0.730 | 0.647 | 0.704 | 0.638 | 0.695 | 0.01% (-6.9 - 6.5) |
| Clinical T Stage | 0.600 | 0.518 | 0.506 | 0.563 | 0.357 | 0.537 | 17.27% (6.2 - 40.4) |
| Clinical N Stage | 0.385 | 0.328 | 0.300 | 0.320 | 0.289 | 0.357 | 17.18% (7.3 - 25.0) |
| Clinical M Stage | 0.252 | 0.070 | 0.048 | 0.065 | 0.043 | 0.090 | 75.01% (64.3 - 83.0) |

Abbreviations: RMSE = root-mean-square error, PFC = proportion of falsely classified entries, Sub = substitution, KNN = k-nearest neighbor, SI = SoftImpute, IS = IterativeSVD, MF = MissForest, DA = denoising autoencoder
<sup>a</sup> Average change refers to the average relative difference between ML imputation methods compared to substitution

### Impact on survival modeling

Cox models incorporating ML imputed clinical data had smaller deviations from models fitted using the complete data compared to substitution. The coefficients estimated using imputed data from MissForest had a MAE of 0.058 and a MSE of 0.011 compared to the “ground truth” Cox model, whereas coefficients estimated using substitution data had the largest MAE of 0.161 and MSE of 0.060. The Cox model C-index was 0.795 when using complete data. Cox models incorporating ML imputed data achieved similar C-indices ranging from 0.787 to 0.801 for all ML methods tested (Table 3).

**Table 3.** Differences in Cox model coefficients and overall model concordance index after incorporation of imputed clinical data.

| **Method** | **MAE** | **MSE** | **C-Index** |
| --- | --- | --- | --- |
| Complete data | - | - | 0.795 |
| Substitution | 0.161 | 0.060 | 0.772 |
| KNN | 0.070 | 0.018 | 0.801 |
| SoftImpute | 0.070 | 0.013 | 0.797 |
| IterativeSVD | 0.099 | 0.023 | 0.787 |
| MissForest | 0.058 | 0.011 | 0.798 |
| Autoencoder | 0.080 | 0.016 | 0.794 |

Abbreviations: MAE = mean absolute error, MSE = mean standard error, C-index = concordance index

### Algorithm runtimes

Substitution had the lowest algorithm runtime, while ML methods required substantially longer runtimes. Substitution required on average 1.54×10^−6^ seconds per patient record. SoftImpute required on average 3.28×10^−4^ seconds per patient record, and MissForest required on average 2.96×10^−3^ seconds per patient record. KNN had the longest runtimes at 3.30×10^−2^ seconds per patient record on average (Figure 4).

## Discussion

Real-world data can be used to generate important insights for cancer patient care. However, missing data remains a substantial issue that will require application of novel imputation strategies. Few studies have specifically compared ML approaches for cancer patients using large scale national registry data. In our study, we demonstrate that ML algorithms can potentially achieve high levels of imputation accuracy while maintaining feature-label relationships in survival modeling. Uniquely, we also show that the performance of ML methods differs by data type and between specific variables of the same type. Among the methods tested, SoftImpute and MissForest had the best performance for continuous and categorical variables. However, while substantial improvement in efficacy was observed with ML methods, these observations did not extend to all variables tested, highlighting the need for further research and variable-specific analysis consideration of ML approaches for missing data correction.

Given clinical information within cancer registries are abstracted from the medical record, missing data may reflect incomplete documentation during the course of routine care. Such data are not missing by chance alone and therefore have substantial implications for patient care and research. Important clinical information is often not readily extracted from the medical record, particularly when structured data fields for a given data element are not available. Even when structured data fields exist, previous research demonstrate that clinical information, such as tumor stage, may be routinely documented as unstructured free text or may not be documented at all.^26,27^ Moreover, while cancer registrars are highly trained in abstracting oncology data, there may be discrepancies in documentation that make reporting challenging.^28^ Given the large number of complex data elements and patients captured in national cancer registries, complete documentation and abstraction of all data is likely an impracticable task. Emerging informatics approaches such as natural language processing and ongoing data standardization efforts have been postulated to increase the speed and completeness of data capture, but recognized challenges remain.^29,30^ Furthermore, missing data is likely not missing completely at random within RWD sources and often introduces systematic differences between patient populations. While methods for handling missing data exist, a recent systematic review of observational time-to-event analysis in oncology showed the majority of studies use complete case analysis.^31,32^ Given the non-random nature of data missingness in real-world settings, this can lead to substantial bias and diminish the generalizability of RWD findings. Therefore, ML approaches for data imputation are also likely to have an increasingly important role in generating clinical insights from RWD sources for cancer patients.

Our study corroborates existing literature showing ML approaches hold substantial promise in missing clinical data imputation within oncology. Previous work examining the application of ML techniques in imputing breast cancer registry data suggested that ML imputed information can be used to improve survival prediction, but the number of patients in the datasets tested were relatively small and the findings may not be generalizable to cancer patients in the United States.^6,33^ It is well recognized that ML methods have variability in their performance among different datasets. Our study corroborates previous work suggesting SoftImpute and MissForest may be highly accurate in imputing clinical data compared to traditional statistical or other ML methods.^34,35^ Our findings are also consistent with studies showing that an increased proportion of missing clinical data likely has an adverse effect on model performance. This is in contrast to earlier results using KNN and singular value decomposition methods on microarray data, which showed no significant deterioration in performance for up to 20% of data missing.^7,16^ This is likely due to loss of inter-feature relationships as a significant portion of the data becomes corrupted. While numerous imputation approaches have been proposed for medical data, few studies have specifically examined their efficacy in real-world cancer registry data.^36–38^ While the recent application of deep learning to missing data imputation has demonstrated performance gains over other ML imputation methods, we did not observe this using the autoencoder model implemented within our study.^19,39,40^ This may indicate that the inter-feature relationships captured within cancer registry data are not well represented within the latent space of deep learning networks, although performance differences are expected with differences in dataset dimensionality and model architecture. The relatively poor ML performance in imputing race within our study also differed from previous work using deep neural networks to impute race/ethnicity using patient disease history, however, a significantly smaller number of features and samples was available in our study.^41^ Nevertheless, our findings reinforce that while ML techniques hold substantial promise in imputing missing clinical data within oncology, data and model specific performances need to be considered.

There are limitations to our study. We examined a single cancer type and registry source, and therefore our results may not be generalizable to other cancer registries. However, our study uniquely uses a large national registry commonly used in observational research and is nevertheless informative for selection of ML approaches in handling missing data. Under our experimental conditions, data was introduced in a missing completely at random fashion. To address this, we also tested a simple scenario of systematic differences in data missingness between metastatic and non-metastatic cancer patients. In real-world scenarios, it is possible that multiple data missingness mechanism exist concurrently including missing at random and missing not at random data. Our study also does not comprehensive test all available ML approaches. Such an undertaking would be impracticable and we have chosen in this study to compare five methods that can be implemented and reproduced with relative ease using open-source libraries. Finally, runtimes were captured are on a high-performance computing server, which may not be reflective of real-world use scenarios where imputation algorithms may be run on local computers with less compute and memory capacity. Nevertheless, the relative performance of each approach is likely similar and will be informative for clinicians and researchers who may wish to incorporate these approaches.

In conclusion, we compared the performance of five ML approaches for imputing missing clinical data among lung cancer patients within a large national registry. Consistent differences in performance were observed by data type and across different thresholds of data missingness, with SoftImpute and MissForest achieving the best performance for continuous and categorical data, respectively. There was substantial variation in the relative performance for each individual variable, however, the overall incorporation of ML imputed clinical data preserved the discriminatory ability of Cox survival models. Taken together, these findings support careful selection of ML methods by data element to improve data completeness for cancer patients.

## Data Availability

All data produced in the present study are available by an application process available to investigators associated with CoC-accredited cancer programs.

## Acknowledgements

This work was funded by a Conquer Cancer Young Investigator Award. Any opinions, findings, and conclusions expressed in this material are those of the author(s) and do not necessarily reflect those of the American Society of Clinical Oncology®, Conquer Cancer®, or the Funder.

This work was presented in part at the American Society of Radiation Oncology Annual Meeting in October 2021.

## Disclosures

**Daniel X. Yang**

Research Funding: ASCO Conquer Cancer, RefleXion Medical

**Yongfeng Hui**

Employment: Amazon (current work completed prior to employment at Amazon)

**Henry S. Park**

Consulting or Advisory Role: AstraZeneca, Galera Medical, Bristol Myers Squib

Research Funding: RefleXion Medical

Travel, Accommodations, Expenses: Bristol Myers Squib

**Sanjay Aneja**

Sanjay Aneja is an Associate Editor for JCO Clinical Cancer Informatics. Journal policy recused the author from having any role in the peer review of this manuscript.

Consulting or Advisory Role: Prophet Consulting (I)

Research Funding: The MedNet, Inc, American Cancer Society, National Science Foundation, Agency for Healthcare Research and Quality, National Cancer Institute, ASCO, Patterson Foundation, Amazon Web Services, RefleXion Medical

Patents, Royalties, Other Intellectual Property: Provisional patent of deep learning optimization algorithm

Travel, Accommodations, Expenses: Prophet Consulting (I), Hope Foundation

Other Relationship: NRG Oncology Digital Health Working Group, SWOG Digital Engagement Committee, ASCO mCODE Technical Review Group

## Figures and Tables

**Supplemental Figure 1.**
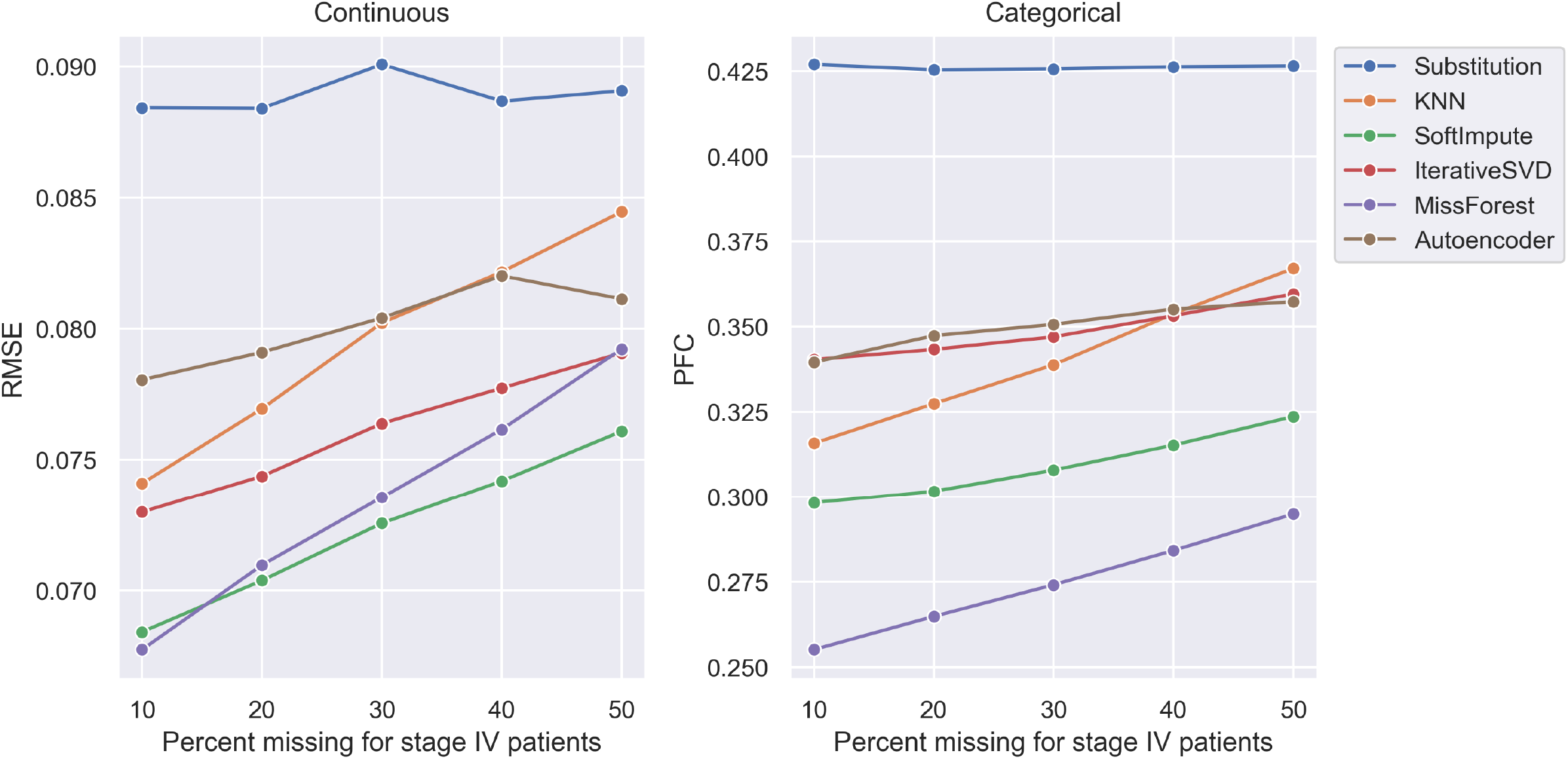
Performance of imputation methods when missingness systematically differed between patients with metastatic versus non-metastatic non-small cell lung cancer. For non-metastatic patients, only 5, 10, 15, 20, and 25 percent of records were spiked with missing data.

**Supplemental Table 1.** National Cancer Database variables used for analysis.

|  |
| --- |
| **Continuous variables** |
| AGE |
| CROWFLY |
| TUMOR_SIZE |
| REGIONAL_NODES_EXAMINED |
| **Categorical variables** |
| SEX |
| INSURANCE_STATUS |
| RX_SUMM_RADIATION |
| RX_SUMM_SURGRAD_SEQ |
| RX_SUMM_CHEMO |
| RX_SUMM_SYSTEMIC_SUR_SEQ |
| RACE |
| RX_SUMM_SURG_PRIM_SITE |
| CDCC_TOTAL_BEST |
| GRADE |
| ANALYTIC_STAGE_GROUP |
| MED_INC_QUAR_16 |
| TNM_CLIN_T |
| TNM_CLIN_N |
| TNM_CLIN_M |

## References

1. Penberthy L, Rivera DR, Ward K. The Contribution of Cancer Surveillance Toward Real World Evidence in Oncology. Semin Radiat Oncol. 2019;29(4):318–322. doi:10.1016/j.semradonc.2019.05.004

2. Booth CM, Karim S, Mackillop WJ. Real-world data: towards achieving the achievable in cancer care. Nat Rev Clin Oncol. 2019;16(5):312–325. doi:10.1038/s41571-019-0167-7

3. Yang DX, Khera R, Miccio JA, et al. Prevalence of Missing Data in the National Cancer Database and Association With Overall Survival. JAMA Netw Open. 2021;4(3):e211793. doi:10.1001/jamanetworkopen.2021.1793

4. Pedersen AB, Mikkelsen EM, Cronin-Fenton D, et al. Missing data and multiple imputation in clinical epidemiological research. Clin Epidemiol. 2017;9:157–166. doi:10.2147/CLEP.S129785

5. Hoskin TL, Boughey JC, Day CN, Habermann EB. Lessons Learned Regarding Missing Clinical Stage in the National Cancer Database. Ann Surg Oncol. 2019;26(3):739–745. doi:10.1245/s10434-018-07128-3

6. Jerez JM, Molina I, García-Laencina Pj, et al. Missing data imputation using statistical and machine learning methods in a real breast cancer problem. Artif Intell Med. 2010;50(2):105–115. doi:10.1016/j.artmed.2010.05.002

7. Beaulieu-Jones BK, Moore JH. MISSING DATA IMPUTATION IN THE ELECTRONIC HEALTH RECORD USING DEEPLY LEARNED AUTOENCODERS. Pac Symp Biocomput. 2017;22:207–218. doi:10.1142/9789813207813_0021

8. Stokes WA, Bronsert MR, Meguid RA, et al. Post-Treatment Mortality After Surgery and Stereotactic Body Radiotherapy for Early-Stage Non-Small-Cell Lung Cancer. J Clin Oncol. 2018;36(7):642–651. doi:10.1200/JCO.2017.75.6536

9. Francis S, Orton A, Stoddard G, et al. Sequencing of Postoperative Radiotherapy and Chemotherapy for Locally Advanced or Incompletely Resected Non-Small-Cell Lung Cancer. J Clin Oncol. 2018;36(4):333–341. doi:10.1200/JCO.2017.74.4771

10. Salazar MC, Rosen JE, Wang Z, et al. Association of Delayed Adjuvant Chemotherapy With Survival After Lung Cancer Surgery. JAMA Oncol. 2017;3(5):610–619. doi:10.1001/jamaoncol.2016.5829

11. Boffa DJ, Rosen JE, Mallin K, et al. Using the National Cancer Database for Outcomes Research: A Review. JAMA Oncol. 2017;3(12):1722–1728. doi:10.1001/jamaoncol.2016.6905

12. Yang CFJ, Chan DY, Speicher PJ, et al. Role of Adjuvant Therapy in a Population-Based Cohort of Patients With Early-Stage Small-Cell Lung Cancer. J Clin Oncol. 2016;34(10):1057–1064. doi:10.1200/JCO.2015.63.8171

13. Robinson CG, Patel AP, Bradley JD, et al. Postoperative radiotherapy for pathologic N2 non-small-cell lung cancer treated with adjuvant chemotherapy: a review of the National Cancer Data Base. J Clin Oncol. 2015;33(8):870–876. doi:10.1200/JCO.2014.58.5380

14. Lin CC, Bruinooge SS, Kirkwood MK, et al. Association Between Geographic Access to Cancer Care, Insurance, and Receipt of Chemotherapy: Geographic Distribution of Oncologists and Travel Distance. J Clin Oncol. 2015;33(28):3177–3185. doi:10.1200/JCO.2015.61.1558

15. Rusthoven CG, Jones BL, Flaig TW, et al. Improved Survival With Prostate Radiation in Addition to Androgen Deprivation Therapy for Men With Newly Diagnosed Metastatic Prostate Cancer. J Clin Oncol. 2016;34(24):2835–2842. doi:10.1200/JCO.2016.67.4788

16. Troyanskaya O, Cantor M, Sherlock G, et al. Missing value estimation methods for DNA microarrays. Bioinformatics. 2001;17(6):520–525. doi:10.1093/bioinformatics/17.6.520

17. Mazumder R, Hastie T, Tibshirani R. Spectral Regularization Algorithms for Learning Large Incomplete Matrices. J Mach Learn Res. 2010;11:2287–2322.

18. Stekhoven DJ, Bühlmann P. MissForest--non-parametric missing value imputation for mixed-type data. Bioinformatics. 2012;28(1):112–118. doi:10.1093/bioinformatics/btr597

19. Lall R, Robinson T. The MIDAS touch: accurate and scalable missing-data imputation with deep learning. Political Analysis. Published online December 14, 2020. Accessed March 2, 2021. https://www.cambridge.org/core/journals/political-analysis

20. Harrell FE, Lee KL, Mark DB. Multivariable prognostic models: issues in developing models, evaluating assumptions and adequacy, and measuring and reducing errors. Stat Med. 1996;15(4):361–387. doi:10.1002/(SICI)1097-0258(19960229)15:4<361::AID-SIM168>3.0.CO;2-4

21. Pedregosa F, Varoquaux G, Gramfort A, et al. Scikit-learn: Machine Learning in Python. Journal of Machine Learning Research. 2011;12(85):2825–2830.

22. Feldman AR Sergey. Fancyimpute: Matrix Completion and Feature Imputation Algorithms. Accessed March 2, 2021. https://github.com/iskandr/fancyimpute

23. Missingpy: Missing Data Imputation for Python. Accessed March 2, 2021. https://github.com/epsilon-machine/missingpy

24. Robinson RL Alex Stenlake, and Thomas. MIDASpy: Multiple Imputation with Denoising Autoencoders. Accessed March 2, 2021. http://github.com/MIDASverse/MIDASpyAneja-Lab-Yale/Aneja-Lab-Public-Impute.

25. Aneja Lab: Yale School of Medicine; 2022. Accessed April 5, 2022. https://github.com/Aneja-Lab-Yale/Aneja-Lab-Public-Impute

26. Evans TL, Gabriel PE, Shulman LN. Cancer Staging in Electronic Health Records: Strategies to Improve Documentation of These Critical Data. J Oncol Pract. 2016;12(2):137–139. doi:10.1200/JOP.2015.007310

27. Sinaiko AD, Barnett ML, Gaye M, Soriano M, Mulvey T, Hochberg E. Association of Peer Comparison Emails With Electronic Health Record Documentation of Cancer Stage by Oncologists. JAMA Netw Open. 2020;3(10):e2015935. doi:10.1001/jamanetworkopen.2020.15935

28. Merriman KW, Broome RG, De Las Pozas G, Landvogt LD, Qi Y, Keating J. Evolution of the Cancer Registrar in the Era of Informatics. JCO Clin Cancer Inform. 2021;5:272–278. doi:10.1200/CCI.20.00123

29. Yim WW, Yetisgen M, Harris WP, Kwan SW. Natural Language Processing in Oncology: A Review. JAMA Oncol. 2016;2(6):797–804. doi:10.1001/jamaoncol.2016.0213

30. Wormeli P, Mazreku J, Pine J, Damesyn M. Next Generation of Central Cancer Registries. JCO Clin Cancer Inform. 2021;5:288–294. doi:10.1200/CCI.20.00177

31. Ibrahim JG, Chu H, Chen MH. Missing data in clinical studies: issues and methods. J Clin Oncol. 2012;30(26):3297–3303. doi:10.1200/JCO.2011.38.7589

32. Carroll OU, Morris TP, Keogh RH. How are missing data in covariates handled in observational time-to-event studies in oncology? A systematic review. BMC Medical Research Methodology. 2020;20(1):134. doi:10.1186/s12874-020-01018-7

33. García-Laencina Pj, Abreu PH, Abreu MH, Afonoso N. Missing data imputation on the 5-year survival prediction of breast cancer patients with unknown discrete values. Comput Biol Med. 2015;59:125–133. doi:10.1016/j.compbiomed.2015.02.006

34. Beaulieu-Jones BK, Lavage DR, Snyder JW, Moore JH, Pendergrass SA, Bauer CR. Characterizing and Managing Missing Structured Data in Electronic Health Records: Data Analysis. JMIR Med Inform. 2018;6(1):e11. doi:10.2196/medinform.8960

35. Waljee AK, Mukherjee A, Singal AG, et al. Comparison of imputation methods for missing laboratory data in medicine. BMJ Open. 2013;3(8). doi:10.1136/bmjopen-2013-002847

36. Howlader N, Noone AM, Yu M, Cronin KA. Use of imputed population-based cancer registry data as a method of accounting for missing information: application to estrogen receptor status for breast cancer. Am J Epidemiol. 2012;176(4):347–356. doi:10.1093/aje/kwr512

37. Cismondi F, Fialho AS, Vieira SM, Reti SR, Sousa JMC, Finkelstein SN. Missing data in medical databases: impute, delete or classify? Artif Intell Med. 2013;58(1):63–72. doi:10.1016/j.artmed.2013.01.003

38. Lin WC, Tsai CF. Missing value imputation: a review and analysis of the literature (2006–2017). Artif Intell Rev. 2020;53(2):1487–1509. doi:10.1007/s10462-019-09709-4

39. Gondara L, Wang K. MIDA: Multiple Imputation Using Denoising Autoencoders. In: Phung D, Tseng VS, Webb GI, Ho B, Ganji M, Rashidi L, eds. Advances in Knowledge Discovery and Data Mining. Lecture Notes in Computer Science. Springer International Publishing; 2018:260–272. doi:10.1007/978-3-319-93040-4_21

40. Phung S, Kumar A, Kim J. A deep learning technique for imputing missing healthcare data. Annu Int Conf IEEE Eng Med Biol Soc. 2019;2019:6513–6516. doi:10.1109/EMBC.2019.8856760

41. Kim JS, Gao X, Rzhetsky A. RIDDLE: Race and ethnicity Imputation from Disease history with Deep LEarning. PLoS Comput Biol. 2018;14(4):e1006106. doi:10.1371/journal.pcbi.1006106

